# Sex-specific impact of body weight on atherosclerotic cardiovascular disease incidence in individuals with and without ideal cardiovascular health

**DOI:** 10.1101/2023.01.16.23284627

**Authors:** Audrey Paulin, Hasanga D. Manikpurage, Jean-Pierre Després, Sébastien Thériault, Benoit J. Arsenault

**Affiliations:** Centre de recherche de l’Institut universitaire de cardiologie et de pneumologie de Québec; VITAM – Centre de recherche en santé durable, Laval University, Quebec (QC), Canada; Department of Kinesiology, Faculty of Medicine, Laval University, Quebec (QC), Canada; Department of Molecular Biology, Medical Biochemistry and Pathology, Faculty of Medicine, Laval University, Quebec (QC), Canada; Department of Medicine, Faculty of Medicine, Laval University, Quebec (QC), Canada

**Keywords:** Body mass index, abdominal adiposity, obesity, cardiovascular disease, lifestyle, metabolic health, ideal cardiovascular health and genetics

## Abstract

**Background:** The impact of an elevated body mass index (BMI) on atherosclerotic cardiovascular diseases (ASCVD) risk in individuals who are “metabolically healthy” is debated. Our objective was to investigate the respective contributions of BMI as well as lifestyle and cardiometabolic risk factors combined to ASCVD incidence in 319,866 UK Biobank participants.

**Methods:** We developed a cardiovascular health score (CVHS) based on four lifestyle and six cardiometabolic parameters. The impact of the CVHS on incident ASCVD (15,699 events) alone and in BMI and waist-to-hip ratio categories was assessed using Cox proportional hazards in women and men separately.

**Results:** In participants with a high CVHS (8-10), those with a BMI ≥ 35.0 kg/m^2^ had a nonsignificant higher ASCVD risk (HR=1.20 [95% CI, 0.84-1.70], p=0.32) compared to those with a BMI of 18.5-24.9 kg/m^2^. In participants with a BMI 18.5-24.9 kg/m^2^, those with a lower CVHS (0-2) had a higher ASCVD risk (HR=4.06 [95% CI, 3.23-5.10], p<0.001) compared to those with a higher CVHS (8-10). When we used the waist-to-hip ratio instead of the BMI, a dose-response relationship between the WHR and ASCVD risk was obtained in healthier participants. Results were similar in women compared to men.

**Conclusions:** In participants of the UK Biobank, the relationship between the BMI and ASCVD incidence in healthy individuals was inconsistent whereas cardiovascular risk factors strongly predicted ASCVD incidence in all BMI categories. Weight inclusive interventions targeting lifestyle-related and metabolic risk factors are likely to prevent cardiovascular outcomes, regardless of their impact on body weight.

## Introduction

Individuals with an elevated body weight are at higher risk of developing many chronic diseases such as atherosclerotic cardiovascular diseases (ASCVD)^1^. Regardless of their body mass index (BMI), individuals with a higher intra-abdominal adipose tissue accumulation and lower peripheral/subcutaneous adipose tissue accumulation also have a higher ASCVD risk. ^2,3^ This is particularly relevevant in men who have a higher ASCVD risk for a given BMI compared to women who tend to accumulate more peripheral that intra-abdominal fat. Several lines of evidence suggest that individuals with an elevated BMI might still be at lower ASCVD risk if they are metabolically healthy (insulin sensitive, normal blood pressure and lipid levels) and/or if they have healthy lifestyle habits (are physically active, do not smoke and eat a healthy diet). ^4,5^ However recent prospective studies have shown that individuals with an elevated BMI might be at higher ASCVD risk, even if they are metabolically healthy^6^ or if they have healthy lifestyle habits^7^. This has led many investigators to conclude that regardless of their metabolic health and lifestyle habits, individuals with an elevated body weight should lose weight to reduce ASCVD risk, even in the absence of conclusive trial evidence showing that intentional weight loss is linked with a lower risk of cardiovascular outcomes and despite the fact that the BMI is not included in ASCVD risk prediction algorithms such as the Pooled Cohort Equations.^8,9^ Many people with suboptimal lifestyle habits may be metabolically healthy and, conversly, many individuals who have healthy lifestyle habits may nevertheless have poor metabolic health.^10^ About 10 years ago, in an effort to improve the health of americans and reduce the public health burden associated with ASCVD, the American Heart Association (AHA) introduced the notion of Life’s Simple 7, a combination of lifestye-related and clinical risk factors that strongly predict ASCVD risk. Individuals with all seven health metrics (not smoking, being physically active, having a healthy dietary pattern and a “healthy weight” as well as controlling blood pressure, blood glucose and cholesterol levels) are considered to have ideal cardiovascular health.^11^ More recently, the AHA update those recommandations to introduce the Life’s essentiels 8, adding sleep duration as a cardiovascular health factor.^12^

To our knowledge, no studies have sought to determine whether BMI predicts ASCVD in individuals with ideal cardiovascular health, that is, in individuals with both healthy lifestyle habits and optimal cardiometabolic health. The potential sex-specific effect of the BMI and body fat distribution patterns on incident ASCVD in healthy individuals is also not well described. The objectives of this study were to investigate the relationship between BMI and incident ASCVD in women and men of the UK Biobank across ideal cardiovascular health categories. In light of studies documenting the association between body fat distribution patterns linked with lower peripheral adiposity and higher abdominal adiposity predicting ASCVD in individuals in any BMI category, we also investigated the impact of the waist-to-hip ratio, a marker of intra-abdominal adipose tissue accumulation in individuals with versus without ideal cardiovascular health.

## Methods

### Study population

The UK Biobank (UKB) is a large-scale prospective cohort study. Over 500,000 participants aged 40-69 years were recruited between 2006 and 2010.^13^ All participants provided written consent at the baseline assessment at one of 22 assessment centres across the United Kingdom. Participants also provided information on lifestyle and health status through questionnaires, interviews, physical measures, blood and urine samples. UKB received approval from the British National Health Service, Northwest-Haydock Research Ethics Committee (16/NW/0274). Data access permission for this study was granted under UKB application 25205. The study samples included 319,866 participants who had a BMI ≥18.5 kg/m^2^, had complete data on lifestyle and metabolic markers and had plasma C-reactive protein (CRP) levels <20 mg/L (as participants with CRP levels ≥20 mg/L might be more likely to have acute rather than systemic inflammation). Participants who had a cardiovascular event before recruitment were excluded. Genetic analyses were performed in 303,110 white British participants with complete genetic/lifestyle information.

### Measurement of body weight and body fat distribution indices

The BMI was obtained at the baseline assessment estimated by impedance measurement or calculated from height and weight measured using the formula weight(kg)/height(m)^2^. Participants were divided into four pre-established BMI groups (18.5-24.9 kg/m^2^, 25-29.9 kg/m^2^, 30-34.9 kg/m^2^, ≥35 kg/m^2^). The waist-to-hip ratio (WHR) was calculated from the baseline measurement of waist circumference and hip circumference using the formula: waist circumference (cm)/hip circumference (cm). Participants were divided in quartiles of WHR for further analysis.

### Cardiovascular health score

We developed a cardiovascular health score (CVHS) based on lifestyle and cardiometabolic health parameters aggregating ten cardiovascular health risk factors. The four lifestyle exposures included smoking status, fruits and vegetables intake, physical activity levels and sleep quality. We defined healthy lifestyle as a smoking status of never or previous smoker, a consumption of five or more daily servings of fruit and vegetable, a minimum of 150 min/week of moderate physical activity or 75 min/week of vigorous physical activity or a combination of moderate and vigorous intensity activity and having a healthy sleep quality. Healthy sleep quality was defined by sleeping 7-9 hours per day, sometimes or never/rarely having insomnia and not having a diagnosis of sleep apnea. Lifestyle information was obtained from the Touchscreen questionnaire recorded at baseline. Cardiometabolic health was assessed by six cardiometabolic health parameters. We defined healthy cardiometabolic health as having a systolic blood pressure (SBP) <130 mmHg and a diastolic blood pressure (DBP) ≤ 80 mmHg without antihypertensive medications, CRP levels <3.0 mg/L, triglyceride levels <2.3 mmol/L, low-density lipoprotein cholesterol (LDL-C) levels <3.0 mmol/L without cholesterol lowering medications, high-density lipoprotein cholesterol (HDL-C) levels >1.0 mmol/L and glycated haemoglobin (HbA_1C_) <42 nmol/mol and no insulin use (except for patients with type 1 diabetes), as previously described for cardiometabolic health.^6^ Participants were then categorised in 11 groups according to both lifestyle and metabolic health parameters.

### Study outcome

The study outcome was incident ASCVD defined as first occurrences or death with the International Classification of Diseases, 10^th^ revision (ICD-10) codes for ischemic stroke (IS; I63) and myocardial infarction (MI; I21-I23). We also added to the ASCVD definition first surgical procedures with Office of Population, Censuses and Surveys: Classification of Interventions and Procedures, version 4 (OPSC-4) for coronary artery bypass grafting (K40.1-40.4, K41.1-41.4, K45.1-45.5), for coronary angioplasty, with or without stenting (K49.1-49.2, K49.8-49.9, K50.2, K75.1-75.4, K75.8-75.9). Dates and causes of hospitalization or death were obtained from death registry, primary care information, hospital admission medical reports and self-report.

### Statistical analyses

Kaplan-Meier analysis were performed to explore the incident ASCVD probabilities in the CVHS categories stratified by sex. Statistical significance through CVHS groups were assessed via log-rank tests. Multivariable Cox proportional hazard models were used to evaluate the association of CVHS categories with the incidence of first ASCVD. Multivariable Cox proportional hazard models were also used to evaluate the respective association of BMI and WHR with the incidence of first ASCVD in CVHS categories. Estimated HRs with their 95% confidence intervals (95% CI) were obtained from those analyses. All analyses were adjusted for age, sex and Townsend deprivation index. Analyses including CVHS categories, BMI and WHR were also adjusted for ethnicity. Dates of recruitment were used as the starting time and participants with event before recruitment were excluded. The end of follow-up was the date of occurrence of the first event (ASCVD), death or censoring. If participant had no event during the follow-up and were still alive, the end of follow-up was the last day of August 2021. The median follow-up was 12.5 years (interquartile range: 11.7-13.2). Schoenfeld tests were performed as a prior to Cox regressions to verify proportionality assumption and Schoenfeld residuals were visually inspected. Harrell’s C-statistic was used to assess the discriminative capacity of incident models and their 95% confidence intervals (CI) were calculated by the bootstrap method (1000 iterations). Differences in C-statistics between models were calculated and 95% confidence intervals (CI) were also obtained by the bootstrap method (1000 iterations). All statistical analyses were conducted using R (v4.1.3).

## Results

The characteristics of study participants at baseline are presented in Table 1. The study sample included 319,866 participants. The percentage of White British individuals varied from 94.0 to 96.9% across groups. These participants were separated according to their CVHS at baseline. Because there were few “very unhealthy” participants to study ASCVD incidence, participants with a score of 0-1 were pooled together. The percentage of males was higher in participants with a lower CVHS. Participants with a lower CVHS also tended to be older, have higher BMI and WHR and a higher social deprivation index. As expected, separation of study participants revealed marked differences in cardiometabolic risk factors across the nine CVHS groups.

**Table 1.** Baseline characteristics of the UK Biobank participants selected for this study by cardiovascular health score categories.

| Factor | Total | CVHS = 10 | CVHS = 9 | CVHS = 8 | CVHS = 7 | CVHS = 6 | CVHS = 5 | CVHS = 4 | CVHS = 3 | CVHS = 2 | CVHS = 0-1 |
| --- | --- | --- | --- | --- | --- | --- | --- | --- | --- | --- | --- |
| N | 319866 | 5010 | 24784 | 67623 | 87141 | 69015 | 39583 | 17997 | 6525 | 1765 | 423 |
| Male* | 141116 (44.1%) | 1073 (21.4%) | 7452 (30.1%) | 26240 (38.8%) | 37319 (42.8%) | 32581 (47.2%) | 20656 (52.2%) | 10281 (57.1%) | 4025 (61.7%) | 1190 (67.4%) | 299 (70.7%) |
| Age, years, mean (SD) | 56.5 (8.1) | 48.8 (7) | 52.6 (8.1) | 55.9 (8.2) | 57.1 (7.9) | 57.5 (7.8) | 57.4 (7.8) | 57.2 (7.8) | 56.6 (7.8) | 56.6 (7.7) | 55.7 (7.7) |
| BMI, kg/m<sup>2</sup>, mean (SD) | 27.4 (4.7) | 23.6 (2.8) | 24.6 (3.2) | 25.7 (3.6) | 26.9 (4.1) | 28.2 (4.6) | 29.6 (5.1) | 30.7 (5.5) | 31.8 (5.8) | 32.9 (6.3) | 33.1 (6.2) |
| Waist-to-hip ratio (Male), cm, mean (SD) | 0.93 (0.06) | 0.87 (0.06) | 0.89 (0.06) | 0.91 (0.06) | 0.93 (0.06) | 0.94 (0.06) | 0.96 (0.06) | 0.97 (0.06) | 0.98 (0.06) | 1.00 (0.07) | 1.00 (0.07) |
| Waist-to-hip ratio (Female), cm, mean (SD) | 0.82 (0.07) | 0.77 (0.05) | 0.78 (0.06) | 0.80 (0.06) | 0.81 (0.06) | 0.83 (0.07) | 0.85 (0.07) | 0.87 (0.07) | 0.89 (0.07) | 0.90 (0.07) | 0.92 (0.06) |
| White ethnicity* | 305379 (95.5%) | 4819 (96.2%) | 23757 (95.9%) | 64991 (96.1%) | 83495 (95.8%) | 65672 (95.2%) | 37504 (94.7%) | 16939 (94.1%) | 6132 (94.0%) | 1669 (94.6%) | 401 (94.8%) |
| Current smoking* | 31451 (9.8%) | 0 (0.0%) | 380 (1.5%) | 2026 (3.0%) | 5251 (6.0%) | 7466 (10.8%) | 7162 (18.1%) | 5119 (28.4%) | 2719 (41.7%) | 982 (55.6%) | 346 (81.8%) |
| Low fruit and vegetable intake* | 58825 (18.4%) | 0 (0%) | 732 (3%) | 3975 (5.9%) | 11028 (12.7%) | 15879 (23%) | 13872 (35%) | 8185 (45.5%) | 3629 (55.6%) | 1168 (66.2%) | 357 (84.4%) |
| Low physical activity* | 138323 (43.2%) | 0 (0%) | 2491 (10.1%) | 12777 (18.9%) | 33744 (38.7%) | 39699 (57.5%) | 27829 (70.3%) | 14191 (78.9%) | 5575 (85.4%) | 1615 (91.5%) | 402 (95%) |
| Poor sleep quality* | 133615 (41.8%) | 0 (0%) | 2553 (10.3%) | 13271 (19.6%) | 34642 (39.8%) | 37547 (54.4%) | 25318 (64%) | 13021 (72.4%) | 5319 (81.5%) | 1547 (87.6%) | 397 (93.9%) |
| Deprivation index, mean (SD) | -1.4 (3.0) | -1.6 (2.9) | -1.6 (2.9) | -1.8 (2.8) | -1.6 (2.9) | -1.4 (3.0) | -1.0 (3.2) | -0.6 (3.4) | -0.1 (3.5) | 0.4 (3.5) | 1.5 (3.6) |
| Systolic BP, mm Hg, mean (SD) | 140 (19.6) | 116.0 (8.4) | 123.9 (15.5) | 136.5 (19.9) | 141.6 (19.1) | 144.0 (18.5) | 145.0 (18.2) | 145.4 (18.0) | 144.7 (17.8) | 144.8 (18.0) | 142.7 (17.6) |
| Diastolic BP, mm Hg, mean (SD) | 82.4 (10.7) | 70.5 (5.9) | 74.4 (8.8) | 80.2 (10.4) | 83.0 (10.3) | 84.5 (10.2) | 85.4 (10.1) | 85.9 (10.3) | 85.7 (10.2) | 85.8 (10.5) | 85.3 (10.2) |
| Antihypertensive medications* | 61397 (19.2) | 0 (0) | 679 (2.7) | 7820 (11.6) | 15526 (17.8) | 16304 (23.6) | 11412 (28.8) | 6195 (34.4) | 2498 (38.3) | 766 (43.4) | 197 (46.6) |
| CRP, mg/L, mean (SD) | 2.6 (4.3) | 0.8 (0.6) | 1.1 (1.7) | 1.4 (2.5) | 2.1 (3.6) | 3 (4.7) | 4.1 (5.3) | 5.1 (6) | 6.1 (6.6) | 7 (6.4) | 8 (5.9) |
| Triglycerides, mmol/L, mean (SD) | 1.7 (1.0) | 0.9 (0.3) | 1.1 (0.4) | 1.3 (0.5) | 1.6 (0.8) | 1.9 (1) | 2.3 (1.2) | 2.7 (1.3) | 3.1 (1.4) | 3.4 (1.4) | 3.7 (1.5) |
| LDL-C, mmol/L, mean (SD) | 3.6 (0.9) | 2.6 (0.3) | 3.1 (0.7) | 3.5 (0.8) | 3.7 (0.8) | 3.7 (0.9) | 3.7 (0.9) | 3.7 (0.9) | 3.6 (0.9) | 3.5 (0.9) | 3.5 (1) |
| HDL-C, mmol/L, mean (SD) | 1.5 (0.4) | 1.6 (0.3) | 1.6 (0.4) | 1.6 (0.4) | 1.5 (0.4) | 1.4 (0.4) | 1.3 (0.3) | 1.2 (0.3) | 1.1 (0.3) | 1.0 (0.2) | 0.9 (0.1) |
| Cholesterol lowering medications* | 48615 (15.2%) | 0 (0.0%) | 572 (2.3%) | 6020 (8.9%) | 11973 (13.7%) | 12583 (18.2%) | 8990 (22.7%) | 5283 (29.4%) | 2245 (34.4%) | 749 (42.4%) | 200 (47.3%) |
| HbA1c, mmol/mol, mean (SD) | 35.9 (6.5) | 32.8 (3.6) | 33.6 (3.6) | 34.4 (3.9) | 35.1 (4.7) | 36.2 (6.1) | 37.8 (7.8) | 40.5 (10.8) | 43.3 (12.5) | 47.4 (14.0) | 49.4 (13.3) |
| On insulin* | 3094 (1.0%) | 17 (0.3%) | 50 (0.2%) | 204 (0.3%) | 464 (0.5%) | 672 (1.0%) | 674 (1.7%) | 564 (3.1%) | 284 (4.4%) | 141 (8.0%) | 24 (5.7%) |

\*N (%)

We investigated the impact of the CVHS on ASCVD incidence. Results presented in Figure 1A revealed a dose-response effect of the CVHS on ASCVD incidence. Compared to participants with a very high CVHS (healthiest, CVHS=10), after adjusting for age, sex, ethnicity and deprivation, those with a very low CVHS (unheatlhiest, CVHS=0-1) had a twelvefold higher ASCVD risk (hazard ratio [HR]=12.58 (95% CI, 8.22-19.25, p<0.001). Because sex differences were identified across CVHS categories (Table 1), sex-specific analyses were performed. Compared to women with a very high CVHS (healthiest, CVHS=10), those with a very low CVHS (unheatlhiest, CVHS=0-1) had a HR for incident ASCVD of 15.80 (95% CI, 7.89-31.80, p<0.001) (Figure 1B). Compared to men with a very high CVHS (healthiest, CVHS=10), those with a very low CVHS (unheatlhiest, CVHS=0-1) had a HR for incident ASCVD of 12.00 (95% CI, 6.68-21.70, p<0.001) (Figure 1C).

**Figure 1.**
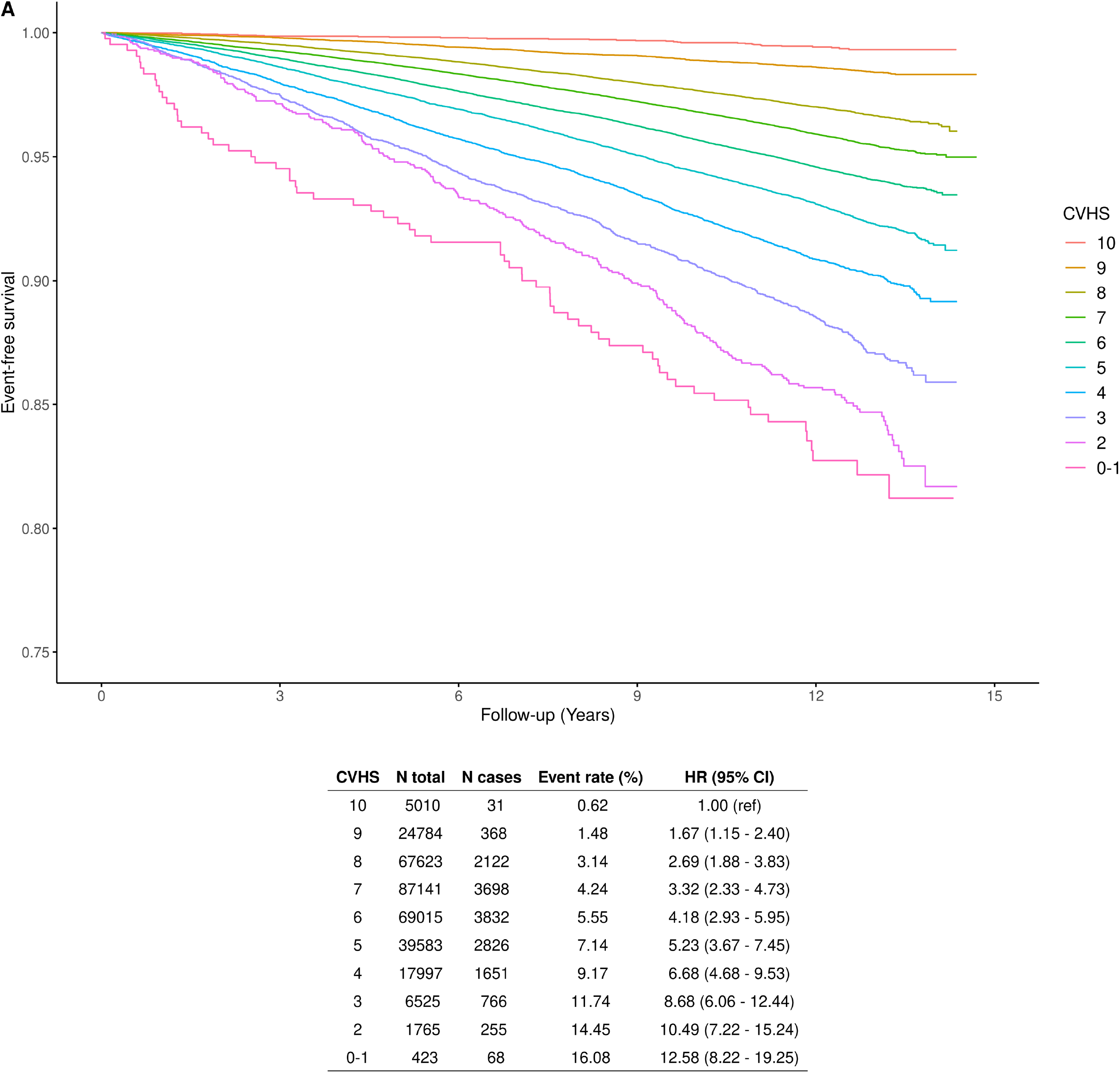
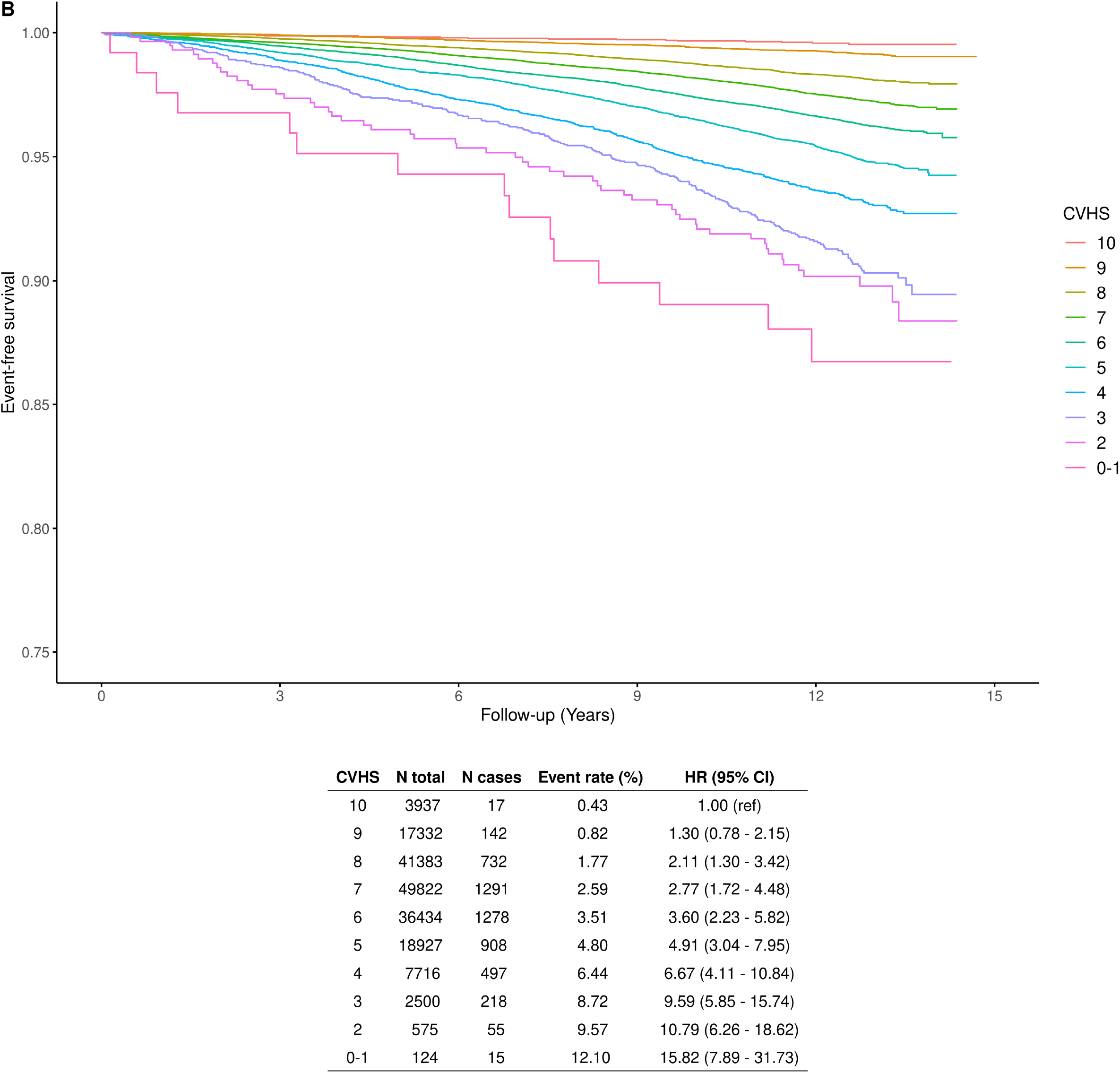
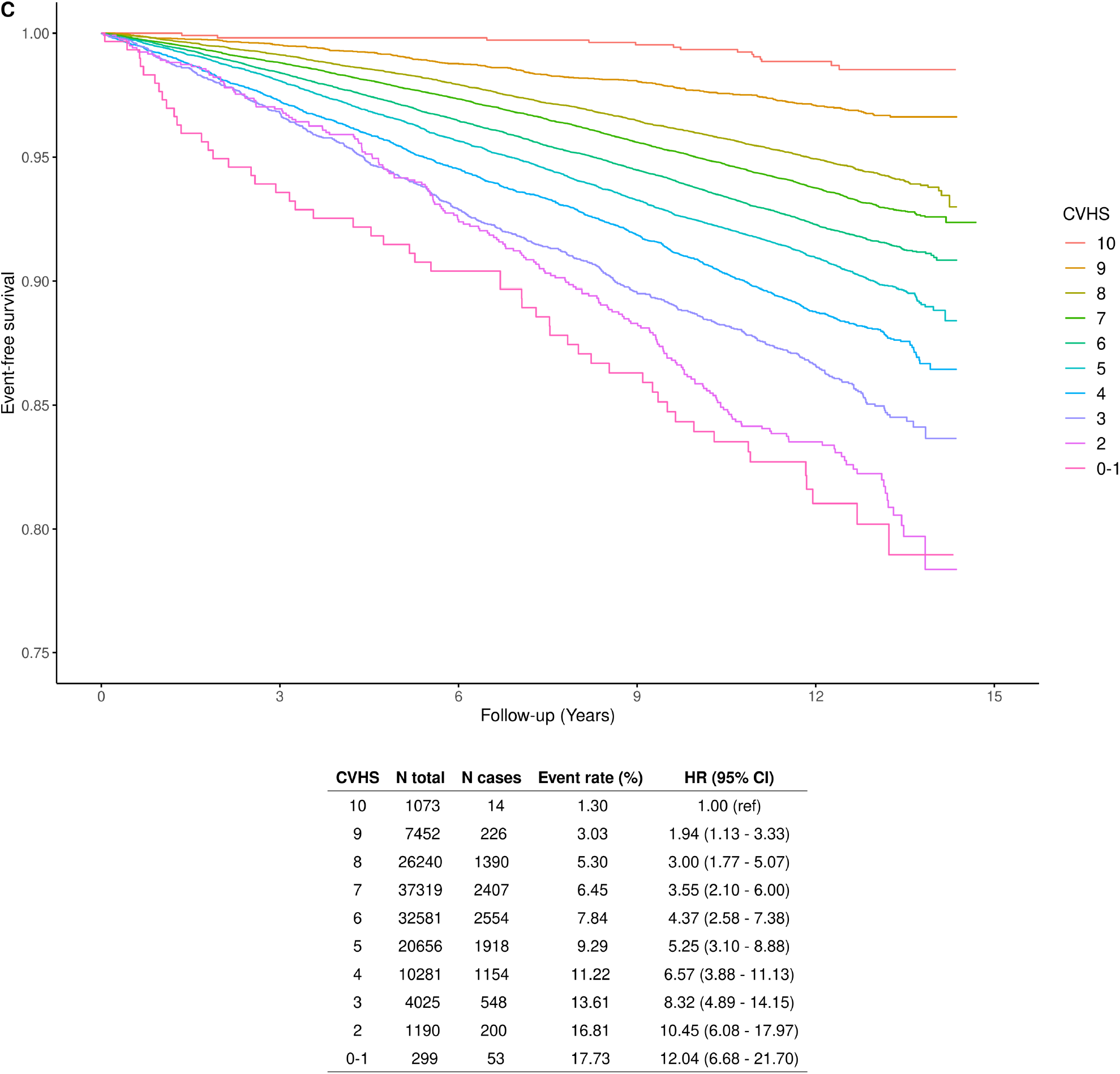
Impact of a cardiovascular health score on atherosclerotic cardiovascular disease incidence in A) all participants of the UK Biobank B) women and C) men. Cox proportional hazards were adjusted for age, ethnicity, deprivation and sex (in the sex-combined analysis). CVHS indicates cardiovascular health score.

The impact of BMI on incident ASCVD in patients in different CVHS categories were investigated. CVHS categories were pooled into four categories to maximise statistical power. In participants with a high CVHS (8-10), although participants with a BMI 25.0-29.9 and 30.0-34.9 kg/m^2^ had a higher ASCVD risk, those with a BMI ≥ 35.0 kg/m^2^ had a nonsignificant slightly higher ASCVD risk (HR=1.20 [95% CI, 0.84-1.70], p=0.32) compared to those with a lower BMI 18.5-24.9 kg/m^2^ (Figure 2). In participants with a BMI 18.5-24.9 kg/m^2^, those with a lower CVHS (0-3) had a higher ASCVD risk (HR=4.06 [95% CI, 3.23-5021], p<0.001) compared to those with a higher CVHS (8-10). Consistent results were observed in both women and men. Altogether, these results suggest that after taking into account lifestyle-related and cardiometabolic risk factors, the relationship between the BMI and incident ASCVD is inconsistent.

**Figure 2.**
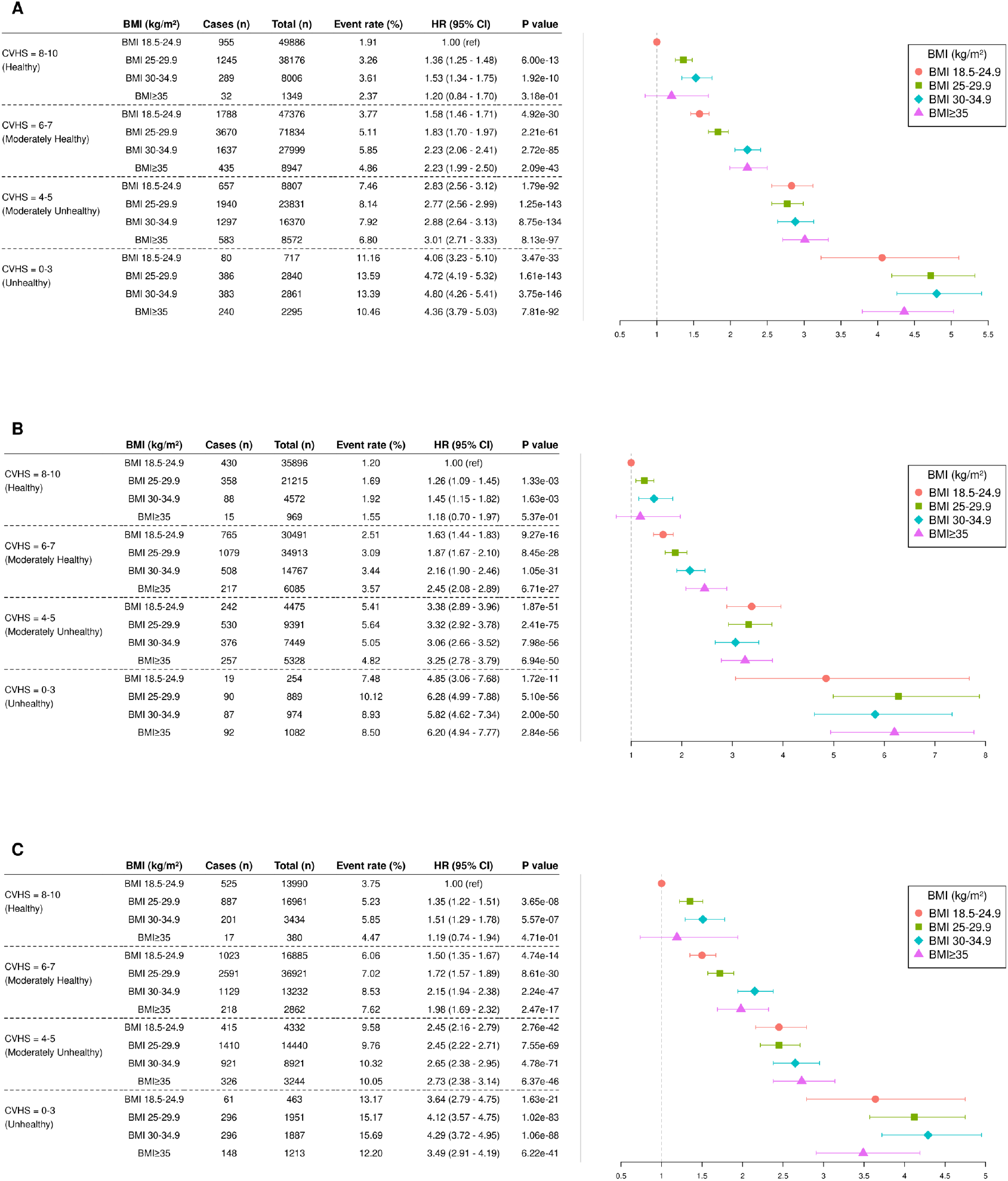
Impact of the body mass index on incident atherosclerotic cardiovascular disease in A) all participants of the UK Biobank B) women and C) men by cardiovascular health score categories. Cox proportional hazards were adjusted for age, ethnicity, deprivation and sex. CVHS indicates cardiovascular health score.

Body fat distribution patterns consistent with higher intra-abdominal and lower subcutaneous/peripheral fat accumulation may be linked with ASCVD, irrespective of the BMI. We investigated the impact of the WHR on incident ASCVD in women and men of the UK Biobank in all BMI categories. Participants were categorized into sex-specific WHR quartiles. Results presented in Figure 3 suggest that in women and men, the WHR is indeed associated with ASCVD. The added predictive value appears to be more important in individuals with lower BMIs. In individuals in the lowest BMI categories (18.5-25.9 kg/m^2^), compared to those in the bottom WHR quartile, individuals in the top WHR quartile had a HR for incident ASCVD of 1.77 (95% CI, 1.58-1.99, p<0.001). These results highlighting the importance of assessing body fat distribution patterns and not only BMI, were similar in women versus men.

**Figure 3.**
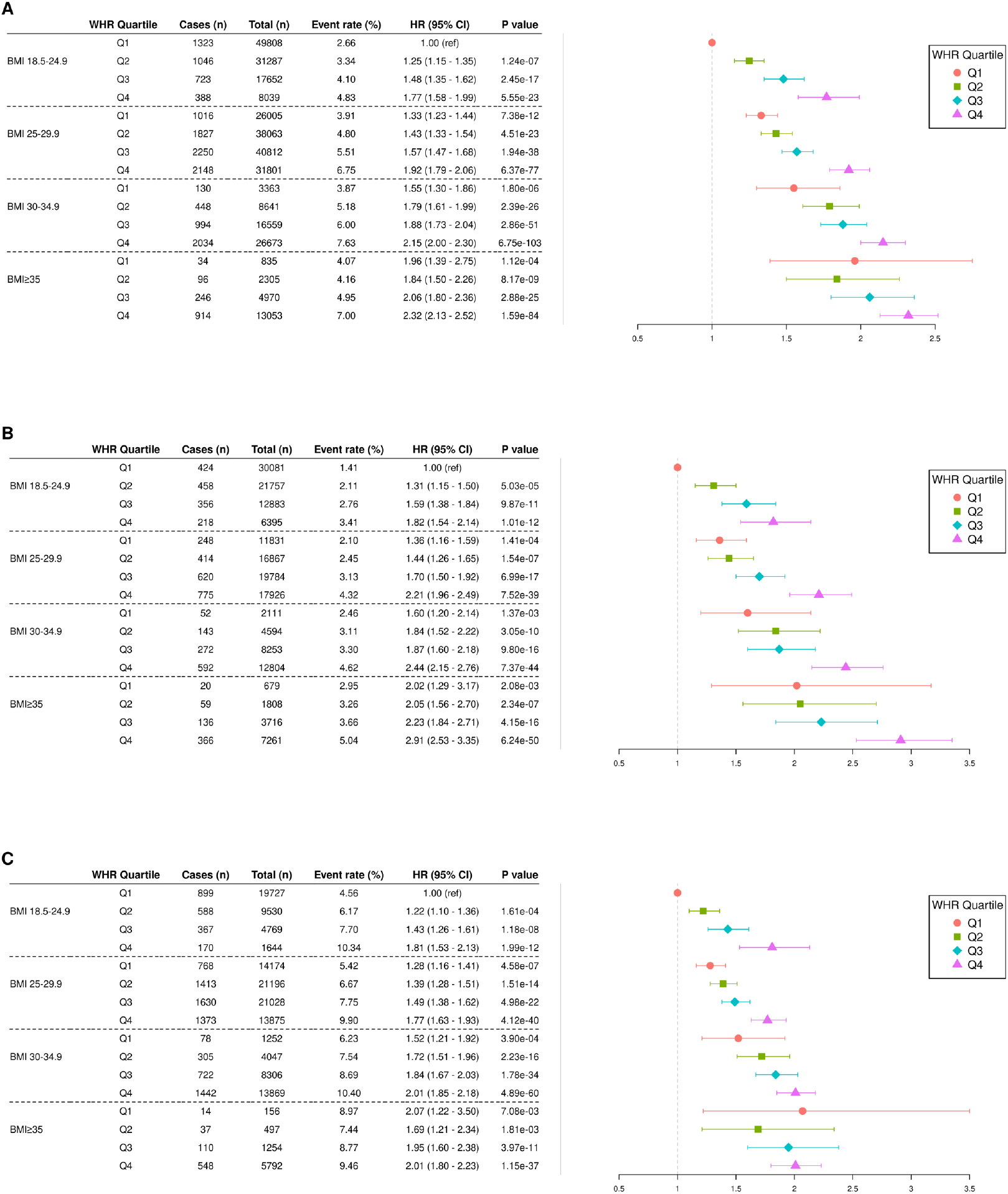
Impact of the waist-to-hip ratio on incident atherosclerotic cardiovascular disease in A) all participants of the UK Biobank B) women and C) men by body mass index categories. Cox proportional hazards were adjusted for age, deprivation and sex. CVHS indicates cardiovascular health score.

In order to investigate whether body fat distribution could be associated with the incidence of ASCVD across CVHS categories, we investigated the impact of the WHR on ASCVD. In participants with a high CVHS (8-10), those with a higher WHR had a higher ASCVD risk (HR=1.57 [95% CI, 1.39-1.77], p<0.001) compared to those with a WHR in the bottom quartile (Figure 4). In participants with a WHR in the bottom quartile, those with a lower CVHS (0-3) had a higher ASCVD risk (HR=4.10 [95% CI, 2.97-5.66], p<0.001) compared to those with a higher CVHS (8-10). These results were similar in women versus men.

**Figure 4.**
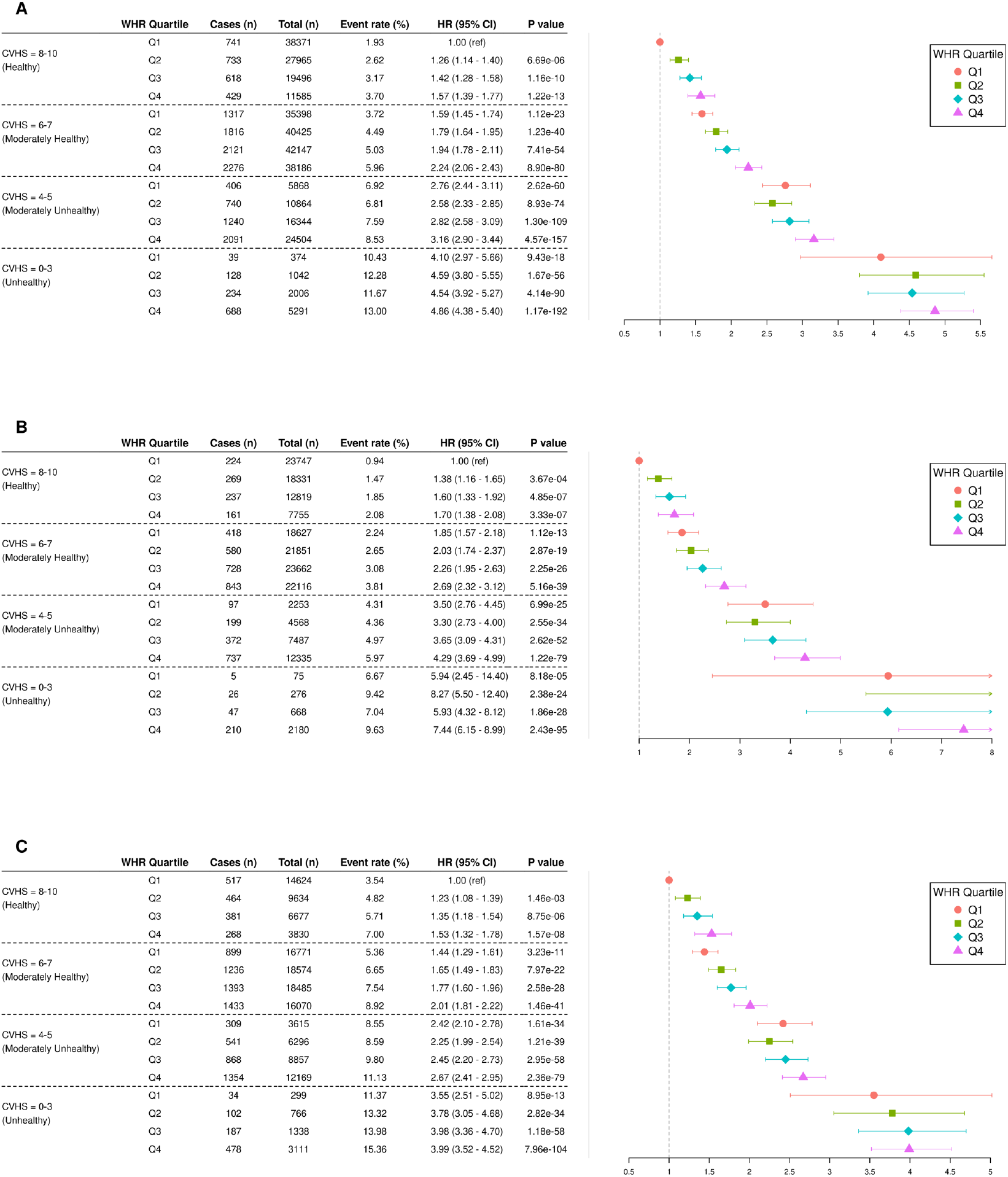
Impact of the waist-to-hip ratio on incident atherosclerotic cardiovascular disease in A) all participants of the UK Biobank B) women and C) men by cardiovascular health score categories. Cox proportional hazards were adjusted for age, ethnicity, deprivation and sex. CVHS indicates cardiovascular health score.

Finally, we investigated whether the BMI and WHR provided added predictive value to that of the CVHS using multivariate Cox regression models. We investigated the association of a model including age, sex and the CVHS and ASCVD incidence as well as two other model that also include BMI and WHR, respectively (Table 2). Adding BMI to the model did not lead to improvements in ASCVD prediction in all participants and in men and only modestly in women. Adding WHR to the model did lead to minor improvements in ASCVD prediction in all participants as well as in women and men separately suggesting that body weight does not lead to better ASCVD prediction when lifestyle and metabolic risk markers are accounted for and that the addition of the WHR could be more clinically useful.

**Table 2.** Comparison of discriminative capacities using Harrell’s C statistic in Cox regression models for the incidence of atherosclerotic cardiovascular diseases adding adiposity indices to the cardiovascular health score (CVHS). Difference between the C-statistic (delta C-statistic) of the studied model (CVHS age and sex) and the C-statistic of the model with or without body mass index or waist to hip ratios are presented.

|  | **Model 1 :**<br>**AGE+SEX** | **Model 2 :**<br>**AGE+SEX+CVHS** | **Model 3 :**<br>**AGE+SEX+CVHS+BMI** | **Difference of C-**<br>**statistic**<br>**Model 2 vs Model 3** | **Model 4 :**<br>**AGE+SEX+CVHS+WHR** | **Difference of C-**<br>**statistic**<br>**Model 2 vs Model 4** |
| --- | --- | --- | --- | --- | --- | --- |
| All | 0.707 (0.703 to 0.711) | 0.731 (0.727 to 0.734) | 0.731 (0.728 to 0.735) | 0.001 (0 - 0.001) | 0.732 (0.729 to 0.736) | 0.002 (0.001 - 0.002) |
| Female | 0.686 (0.679 to 0.693) | 0.723 (0.717 to 0.729) | 0.723 (0.694 to 0.701) | 0.001 (0 - 0.001) | 0.726 (0.718 to 0.725) | 0.003 (0.002 - 0.004) |
| Male | 0.642 (0.636 to 0.646) | 0.67 (0.665 to 0.675) | 0.671 (0.694 to 0.701) | 0.001 (0.001 - 0.002) | 0.672 (0.718 to 0.725) | 0.002 (0.001 - 0.003) |

## Discussion

In this large sample of participants included in the UK Biobank, a CVHS including both lifestyle-related and cardiometabolic risk factors was strongly and dose-dependently associated with ASCVD incidence. This observation was consistent across BMI categories. Participants with the lowest CVHS score had a twelvefold increased ASCVD risk compared to those with the highest CVHS. While individuals with a lower CVHS had, on average higher adiposity indices, we found that the relationship between the BMI and incident ASCVD was inconsistent, especially when compared to the CVHS which showed dose-dependent effect on ASCVD incidence. The WHR, a marker of intra-abdominal adipose tissue accumulation/low peripheral adipose tissue accumulation, was however associated with ASCVD in participants with a high CVHS. These results, which were comparable when women and men were investigated separately, suggest that people with an elevated BMI could be at low ASCVD risk if they have healthy lifestyle habits and are able to control cardiometabolic risk factors and that people with a low CVHS could be at very high ASCVD risk even if they have a “normal” BMI.

Our findings underscore the importance of maintaining healthy lifestyle habits as well as controlling cardiometabolic risk factors, regardless of the impact of these interventions on body weight. Several studies have shown that the adoption of healthier lifestyle habits reduces ASCVD incidence, even if these interventions do not influence body weight. For instance, randomized clinical trials of patients with and without ASCVD revealed that the adoption of healthy dietary patterns such as the Mediterranean diet significantly reduces ASCVD.^14-16^ Additionally, smoking cessation has a major incidence on ASCVD risk reduction, despite being linked with weight gain, the risk associated with weight gain being trivial compared to the benefits of smoking cessation.^17^ Controlling cholesterol levels, blood pressure and even inflammation reduces ASCVD events in the absence of weight changes.^18-20^ Some drugs used for glucose control such as thiazolidinediones (TZD) also provide cardiovascular benefits, despite being associated with weight gain.^21^ Interestingly, weight gain associated with the TZD pioglitazone seems to be limited to peripheral or subcutaneous body fat depots.^22^ This finding is in line with the results of the current study which shows a non-consistent effect of BMI on ASCVD risk in individuals with ideal cardiovascular health. However, participants with an elevated WHR are at higher ASCVD risk, even if they have ideal cardiometabolic health. In this regard, interventions trials documenting the impact of physical activity on health outcomes revealed that despite being associated with minimal weight loss, physical activity may promote the mobilization of intra-abdominal and ectopic lipid deposition such as liver fat and improve cardiometabolic health.^23^ Altogether, these studies support the notion that “metabolically healthy obese” individuals may be at lower ASCVD risk because they have lower intra-abdominal adipose tissue accumulation and/or higher peripheral or subcutaneous adipose tissue accumulation^24^ or higher lean body mass^25,26^.

Results of the current study provide additional support that lifestyle and weight inclusive approaches (approaches that target health and lifestyle-related factors regardless of their impact on body weight) should be preferred over weight-centric approaches for the prevention of ASCVD. In contrast to lifestyle and weight inclusive approaches, weight-centric approaches for the prevention of ASCVD have been shown to be largely ineffective.^8,9,27^ Increasing evidence also suggest that weight-centric approaches are not sustainable for most people and may even be harmful, for instance by contributing to weight stigma.^28^ Weight stigma often results in emotional distress (body dissatisfaction, low self-esteem and well-being, depression, etc.) and psychological stress, all of which are risk factors for ASCVD.^29,30^ Weight-centric approaches also often have the unintended consequence of setting up patients for lifelong weight cycling patterns. Constant dieting and weight cycling are associated with a higher risk of developing eating disorders, chronic stress, poor cardiometabolic and mental health and a higher risk of long-term cardiovascular events.^31,32^ Our results also reinforce the notion that measuring both cardiometabolic and lifestyle-related risk factors is primary care setting might be clinically useful. Optimal assessment of these risk factors, which are relatively easy to measure, might identify individuals at high ASCVD risk (despite having low body weight) through the identification of actionable ASCVD risk factors.

Strengths of the current study include the large sample size from a contemporary cohort that includes both men and women. We also used different classification and definitions of total and regional adiposity, which reinforced the notion that body fat localization may be an important determinant of cardiovascular health. On the other hand, it must be noted that the study population was predominantly white and slightly healthier than the general population. The predictive value of total and regional adiposity across cardiovascular health categories should be investigated in different populations with different levels of risk and different patterns of body fat distributions. We also only investigated the impact of the CVHS on ASCVD and not on other cardiovascular outcomes (heart failure, peripheral artry disease, atrial fibrillation, etc.) due to a lower number of events. Finally, this prospective study was observational by design and causality cannot be infered. Bias due to unmeasured confounders and reverse causality cannot be excluded.

Our study results are comparable to an analysis of the NHANES Survey III showing that BMI was not associated with all-cause mortality in patients with healthy lifestyle parameters^5^ and with our previous investigation of the EPIC-Norfolk study showing that people with an elevated body weight were not at higher coronary heart disease (CHD) risk if they were metabolically healthy and had low CRP levels^4^. In a large analysis of 3.5 million participants from the United Kingdom followed for 5.4 years, Caleyachetty et al. reported that individuals with an elevated BMI (>30 kg/m^2^) were at higher CHD risk compared to those with a BMI 18.5-24.9 kg/m^2^, even if they did not have metabolic abnormalities (defined by the presence of diabetes, hypertension and hyperlipidemia).^33^ In that study, cardiometabolic risk factors were not measured and lifestyle-related factors were not obtained. In a previous analysis of the UK Biobank, Zhou et al. reported that individuals with an elevated BMI (>30 kg/m^2^) were at higher ASCVD risk compared to those with a BMI <30 kg/m^2^, even if they did not have metabolic abnormalities (defined by same criteria as the present study).^6^ However, none of these studies included lifestyle-related parameters in their definition of metabolic health. The previous analysis of the UK Biobank also only investigated participants with a BMI higher or lower than 30 kg/m^2^. Another analysis of the UK Biobank investigated the predictive value of BMI in patients separated into metabolic health (defined by the presence of diabetes, hypertension and hyperlipidemia) or lifestyle-related factors but not in a comprehensive CVHS including both lifestyle and cardiometabolic risk such as the one used in the current study (and as recommended by the AHA). Investigators from these studies concluded that regardless of their lifestyle habits or cardiometabolic health, individuals living in larger bodies should lose weight to decrease their ASCVD risk. By showing that the BMI was an inconsistent predictor of ASCVD in people with a high CVHS, our results do not provide support for this conclusion. Results of our analysis suggest that measuring and monitoring actionnable lifestyle-related risk and cardiometabolic risk factors is crucial to prevent ASCVD events. In conclusion, we propose that weight inclusive interventions targeting lifestyle-related, metabolic risk factors and intra-abdominal fat accumulation, rather than weight-centric approaches are likely to improve the cardiovascular health and overall well-being of people of all body sizes.

## Data Availability

The individual-level data can be obtained via the UK Biobank.

## Acknowledgements

We would like to thank all study participants and staff of the UK Biobank. HDM holds a doctoral research award from the Quebec Heart and Lung Institute. BJA holds a senior scholar award from the Fonds de recherche du Québec: Santé and is supported by grants from the Canadian Institutes of Health and the Foundation of the Quebec Heart and Lung Institute.

## References

1. Pischon T, Boeing H, Hoffmann K, Bergmann M, Schulze MB, Overvad K, van der Schouw YT, Spencer E, Moons KGM, Tjonneland A, et al. General and Abdominal Adiposity and Risk of Death in Europe. New England Journal of Medicine. 2008;359:2105–2120. doi: 10.1056/NEJMoa0801891

2. Canoy D, Boekholdt SM, Wareham N, Luben R, Welch A, Bingham S, Buchan I, Day N, Khaw KT. Body fat distribution and risk of coronary heart disease in men and women in the European prospective investigation into cancer and nutrition in Norfolk cohort - A population-based prospective study. Circulation. 2007;116:2933–2943. doi: 10.1161/circulationaha.106.673756

3. Neeland IJ, Ross R, Despres JP, Matsuzawa Y, Yamashita S, Shai I, Seidell J, Magni P, Santos RD, Arsenault B, et al. Visceral and ectopic fat, atherosclerosis, and cardiometabolic disease: a position statement. Lancet Diabetes & Endocrinology. 2019;7:715–725. doi: 10.1016/s2213-8587(19)30084-1

4. Van Wijk DF, Boekholdt SM, Arsenault BJ, Ahmadi-Abhari S, Wareham NJ, Stroes ESG, Khaw KT. C-Reactive Protein Identifies Low-Risk Metabolically Healthy Obese Persons: The European Prospective Investigation of Cancer-Norfolk Prospective Population Study. Journal of the American Heart Association. 2016;5. doi: 10.1161/jaha.115.002823

5. Matheson EM, King DE, Everett CJ. Healthy Lifestyle Habits and Mortality in Overweight and Obese Individuals. Journal of the American Board of Family Medicine. 2012;25:9–15. doi: 10.3122/jabfm.2012.01.110164

6. Zhou ZY, Macpherson J, Gray SR, Gill JMR, Welsh P, Celis-Morales C, Sattar N, Pell JP, Ho FK. Are people with metabolically healthy obesity really healthy? A prospective cohort study of 381,363 UK Biobank participants. Diabetologia. 2021;64:1963–1972. doi: 10.1007/s00125-021-05484-6

7. Heath L, Jebb SA, Aveyard P, Piernas C. Obesity, metabolic risk and adherence to healthy lifestyle behaviours: prospective cohort study in the UK Biobank. Bmc Medicine. 2022;20. doi: 10.1186/s12916-022-02236-0

8. Powell-Wiley TM, Poirier P, Burke LE, Despres JP, Gordon-Larsen P, Lavie CJ, Lear SA, Ndumele CE, Neeland IJ, Sanders P, et al. Obesity and Cardiovascular Disease: A Scientific Statement From the American Heart Association. Circulation. 2021;143:E984–E1010. doi: 10.1161/cir.0000000000000973

9. Wing R, Bolin P, Brancati FL, Bray GA, Clark JM, Coday M, Crow RS, Curtis JM, Egan CM, Espeland MA, et al. Cardiovascular Effects of Intensive Lifestyle Intervention in Type 2 Diabetes. New England Journal of Medicine. 2013;369:145–154. doi: 10.1056/NEJMoa1212914

10. Folsom AR, Yatsuya H, Nettleton JA, Lutsey PL, Cushman M, Rosamond WD, Investigators AS. Community Prevalence of Ideal Cardiovascular Health, by the American Heart Association Definition, and Relationship With Cardiovascular Disease Incidence. Journal of the American College of Cardiology. 2011;57:1690–1696. doi: 10.1016/j.jacc.2010.11.041

11. Spring B, Ockene JK, Gidding SS, Mozaffarian D, Moore S, Rosal MC, Brown MD, Vafiadis DK, Cohen DL, Burke LE, et al. Better Population Health Through Behavior Change in Adults A Call to Action. Circulation. 2013;128:2169–2176. doi: 10.1161/01.cir.0000435173.25936.e1

12. Lloyd-Jones DM, Allen NB, Anderson CAM, Black T, Brewer LC, Foraker RE, Grandner MA, Lavretsky H, Perak AM, Sharma G, et al. Life’s Essential 8: Updating and Enhancing the American Heart Association’s Construct of Cardiovascular Health: A Presidential Advisory From the American Heart Association. Circulation. 2022;146:e18–e43. doi: 10.1161/cir.0000000000001078

13. Sudlow C, Gallacher J, Allen N, Beral V, Burton P, Danesh J, Downey P, Elliott P, Green J, Landray M, et al. UK Biobank: An Open Access Resource for Identifying the Causes of a Wide Range of Complex Diseases of Middle and Old Age. Plos Medicine. 2015;12. doi: 10.1371/journal.pmed.1001779

14. de Lorgeril M, Salen P, Martin JL, Monjaud I, Delaye J, Mamelle N. Mediterranean diet, traditional risk factors, and the rate of cardiovascular complications after myocardial infarction - Final report of the Lyon Diet Heart Study. Circulation. 1999;99:779–785. doi: 10.1161/01.cir.99.6.779

15. Estruch R, Ros E, Salas-Salvado J, Covas MI, Corella D, Aros F, Gomez-Gracia E, Ruiz-Gutierrez V, Fiol M, Lapetra J, et al. Primary Prevention of Cardiovascular Disease with a Mediterranean Diet Supplemented with Extra-Virgin Olive Oil or Nuts. New England Journal of Medicine. 2018;378. doi: 10.1056/NEJMoa1800389

16. Delgado-Lista J, Alcala-Diaz JF, Torres-Pena JD, Quintana-Navarro GM, Fuentes F, Garcia-Rios A, Ortiz-Morales AM, Gonzalez-Requero AI, Perez-Caballero AI, Yubero-Serrano EM, et al. Long-term secondary prevention of cardiovascular disease with a Mediterranean diet and a low-fat diet (CORDIOPREV) a randomised controlled trial. Lancet. 2022;399:1876–1885. doi: 10.1016/s0140-6736(22)00122-2

17. Sahle BW, Chen W, Rawal LB, Renzaho AMN. Weight Gain After Smoking Cessation and Risk of Major Chronic Diseases and Mortality. Jama Network Open. 2021;4. doi: 10.1001/jamanetworkopen.2021.7044

18. Collins R, Reith C, Emberson J, Armitage J, Baigent C, Blackwell L, Blumenthal R, Danesh J, Smith GD, DeMets D, et al. Interpretation of the evidence for the efficacy and safety of statin therapy. Lancet. 2016;388:2532–2561. doi: 10.1016/s0140-6736(16)31357-5

19. Dahlof B, Sever PS, Poulter NR, Wedel H, Beevers DG, Caulfield M, Collins R, Kjeldsen SE, Kristinsson A, McInnes GT, et al. Prevention of cardiovascular events with an antihypertensive regimen of amlodipine adding perindopril as required versus atenolol adding bendroflumethiazide as required, in the Anglo-Scandinavian Cardiac Outcomes Trial-Blood Pressure Lowering Arm (ASCOT-BPLA): a multicentre randomised controlled trial. Lancet. 2005;366:895–906. doi: 10.1016/s0140-6736(05)67185-1

20. Ridker PM, Everett BM, Thuren T, MacFadyen JG, Chang WH, Ballantyne C, Fonseca F, Nicolau J, Koenig W, Anker SD, et al. Antiinflammatory Therapy with Canakinumab for Atherosclerotic Disease. New England Journal of Medicine. 2017;377:1119–1131. doi: 10.1056/NEJMoa1707914

21. de Jong M, van der Worp HB, van der Graaf Y, Visseren FLJ, Westerink J. Pioglitazone and the secondary prevention of cardiovascular disease. A meta-analysis of randomized-controlled trials. Cardiovascular Diabetology. 2017;16. doi: 10.1186/s12933-017-0617-4

22. Bertrand OF, Poirier P, Rodes-Cabau J, Rinfret S, Title LM, Dzavik V, Natarajan M, Angel J, Batalla N, Almeras N, et al. Cardiometabolic effects of rosiglitazone in patients with type 2 diabetes and coronary artery bypass grafts: A randomized placebo-controlled clinical trial. Atherosclerosis. 2010;211:565–573. doi: 10.1016/j.atherosclerosis.2010.06.005

23. Cowan TE, Brennan AM, Stotz PJ, Clarke J, Lamarche B, Ross R. Separate Effects of Exercise Amount and Intensity on Adipose Tissue and Skeletal Muscle Mass in Adults with Abdominal Obesity. Obesity. 2018;26:1696–1703. doi: 10.1002/oby.22304

24. Arsenault BJ, Lachance D, Lemieux I, Almeras N, Tremblay A, Bouchard C, Perusse L, Despres JP. Visceral adipose tissue accumulation, cardiorespiratory fitness, and features of the metabolic syndrome. Archives of Internal Medicine. 2007;167:1518–1525. doi: 10.1001/archinte.167.14.1518

25. Baker JF, Harris T, Rapoport A, Ziolkowski SL, Leonard MB, Long J, Zemel B, Weber DR. Validation of a description of sarcopenic obesity defined as excess adiposity and low lean mass relative to adiposity. Journal of Cachexia Sarcopenia and Muscle. 2020;11:1580–1589. doi: 10.1002/jcsm.12613

26. Despres JP. Taking a closer look at metabolically healthy obesity COMMENT. Nature Reviews Endocrinology. 2022;18:131–132. doi: 10.1038/s41574-021-00619-6

27. Gaesser GA, Angadi SS. Obesity treatment: Weight loss versus increasing fitness and physical activity for reducing health risks. Iscience. 2021;24. doi: 10.1016/j.isci.2021.102995

28. Mauldin K, May M, Clifford D. The consequences of a weight-centric approach to healthcare: A case for a paradigm shift in how clinicians address body weight. Nutrition in Clinical Practice. doi: 10.1002/ncp.10885

29. Rukh G, Ahmad S, Lind L, Schioth HB. Evidence of a Causal Link Between the Well-Being Spectrum and the Risk of Myocardial Infarction: A Mendelian Randomization Study. Frontiers in Genetics. 2022;13. doi: 10.3389/fgene.2022.842223

30. Tomiyama AJ, Carr D, Granberg EM, Major B, Robinson E, Sutin AR, Brewis A. How and why weight stigma drives the obesity ‘epidemic’ and harms health. Bmc Medicine. 2018;16. doi: 10.1186/s12916-018-1116-5

31. Tomiyama AJ. Weight stigma is stressful. A review of evidence for the Cyclic Obesity/Weight-Based Stigma model. Appetite. 2014;82:8–15. doi: 10.1016/j.appet.2014.06.108

32. Bangalore S, Fayyad R, Laskey R, DeMicco DA, Messerli FH, Waters DD. Body-Weight Fluctuations and Outcomes in Coronary Disease. New England Journal of Medicine. 2017;376:1332–1340. doi: 10.1056/NEJMoa1606148

33. Caleyachetty R, Thomas GN, Toulis KA, Mohammed N, Gokhale KM, Balachandran K, Nirantharakumar K. Metabolically Healthy Obese and Incident Cardiovascular Disease Events Among 3.5 Million Men and Women. Journal of the American College of Cardiology. 2017;70:1429–1437. doi: 10.1016/j.jacc.2017.07.763

